# Does pre-notification increase questionnaire response rates: a nested randomised control trial

**DOI:** 10.1101/2021.02.19.21252107

**Authors:** Benjamin Woolf, Phil Edwards

**Author notes:** **Named Contact and Address:** Benjamin Woolf; University of Bristol, Department of Psychological Science, 5 Priory Road.

## Abstract

**Background:** Study results can be badly affected by non-response. One way to potentially reduce non-response is by sending potential study participants advance communication. During the update of a systematic review examining the effect of pre-notification on response rates, a number of study authors needed to be contacted for further information.

**Objectives:** To conduct an RCT to investigate the effect of pre-notification, nested within the request for further information for a systematic review.

**Methods:** Study authors included in the systematic review, whose studies were at unclear risk of bias, and who were contactable, were randomly sent or not set a pre-notification email prior to being sent the request for further information email.

**Results:** At the end of follow up, 14/33 (42.4%) authors in the pre-notification condition had returned responses to the questionnaire, and 18/42 (42.9%). There was not evidence of a difference between these groups.

**Conclusions:** This study’s results do not support the hypothesis that pre-notification does increase response from participants.

## Introduction

Loss to follow-up and non-response are undesirable features to have in a study. Loss to follow up reduces study power by reducing the number of participants on which data is collected for and increase cost through wasted resources. More worryingly, it also introduces risk of selection bias, and therefore potentially perturbs a studies internal validity, and increase study costs [1,2].

It is therefore important to find ethical ways of reducing non-response. One potential method for doing so is notifying participants of the attempt to send collect data in advance. When questionnaires are the mode of data collection this is often termed ‘pre-notification’ or ‘pre-contact’. In 2009, Edwards et al. published a systematic review of randomised control trials evaluating methods of reducing questionnaire non-response. They found that pre-contact increased response when compared to no pre-contact (OR = 1.5, 95% CI 1.26-1.78, for response after first questionnaire administration, and OR = 1.45, 95% CI 1.29-1.63 for response after final questionnaire administration) [3]. However, this study is now a decade old, so we started an update of this systematic review [4].

A large proportion of studies did not provide enough information for an unambiguous risk of bias evaluation using the Cochrane Risk of Bias tool. This study is nested within the author follow up of the updated aforementioned systematic review, and aimed to provide further evidence on the question of whether pre-notification increases response rates to questionnaires.

## Methods

### Trial design

This study is a two arm randomised trial, with participants randomised with a 50% chance to the intervention (pre-notification) and control (no pre-notification) arm.

### Participants

Participants were eligible to be entered into the study if they were the corresponding author a study deemed eligible for a systematic review into the effect of pre-notification on response rates, but had provided insufficient detail in the written report for the paper to be judged as high or low risk of bias. In cases in which valid contact details for the corresponding authors were not accessible, other study authors were included in the study instead.

Participants were excluded if no means of email communication was found. This was established primarily by checking the stated address in papers. The validity of the address was confirmed by checking author’s university/personal web-page. In cases of discrepancy the emails were sent to both accounts. If no email could be found Research Gate were checked as another possible means of contact. No power analysis was therefore conducted to determine the sample size required for this study.

### Interventions

After randomisation those participants allocated to the intervention arm received a pre-notification email. One day later, they were emailed the questionnaire. Follow up contacts were sent at one, and two weeks after the initial sending. Participants in the control group received the same regimen, except that they did not receive the pre-notification. Other than the pre-notification, all communication to the two arms were sent on the same day. The pre-written communication and questionnaire are displayed bellow.

### Outcomes

The primary outcome in this paper is the final response rate. This is defined by the number of responses at two weeks after the sending of all follow up communication divided by the total number of included participants. The secondary outcome is the response rate prior to follow up. The second outcome is defined as the number of responses before the sending of follow up communication divided by the total number of included participants.

### Randomisation

Sequence generation: the intervention and control arm were assigned numerical values (1 or 0 respectively). The sequence was then generated using the random number generator on a Casio fx-85GT PLUS calculator.

Allocation concealment: Prior to allocation, study authors were psudo–anonymised by physically masking identifiable details.

Blinding of personnel and participants: No active blinding of participants or personnel occurred. However, no material risk of bias should be introduced. Because participants were unaware of having been randomised, any effect of treatment could not be due to the effect of knowing that they had been specially selected for an intervention which others had not got. Although the participant still knew they had received the pre-notification, this knowledge is part of the effect of a pre-notification.

Likewise, although unblidned, because all communication was pre-written, study personnel do not have the ability to influence the experience or perceptions of potential participants, as their only means of communication with each other is through a pre-written pro-forma message.

Blinding of outcome assessment: The number of questionnaires in each arm returned was logged. Data was pseudo anonymised prior to statistical analyse.

### Statistical methods

Results will be computed by calculating the proportion of responses in allotted times, the difference in these proportions, with corresponding 95% confidence intervals, and a t-test.

## Results

### Participant flow

See Figure 1

**Figure 1:**
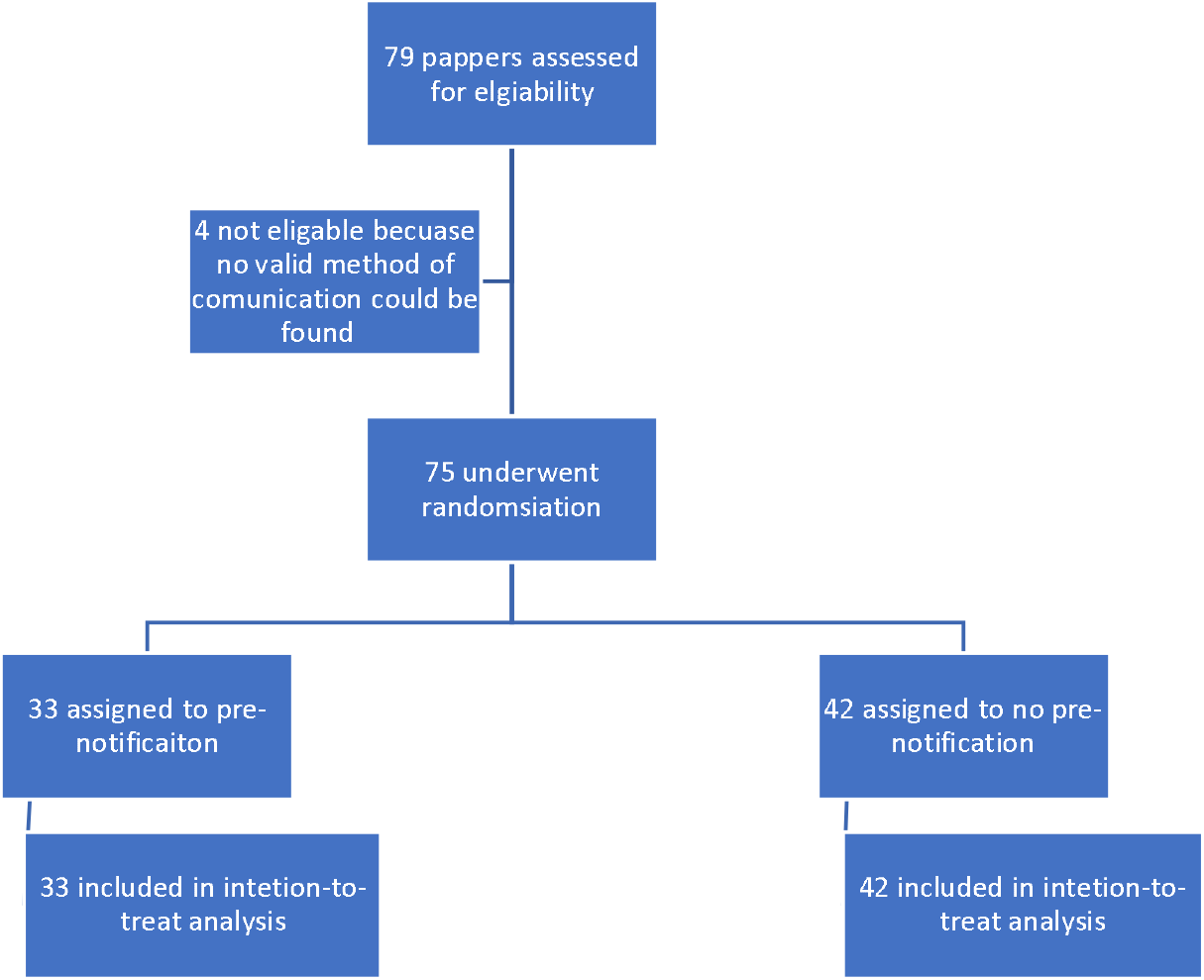
flow diagram of participant recruitment

### Recruitment

Participants were recruited into this trial implicitly through the request for information. These were sent in early June 2019. As defined in the methods section, participants were then given a month for follow-up. The trail was stopped at the end of this time period. 8 authors were contacted through Research Gate, with an equal split across arms.

### Numbers analysed

33 studies are included in the analysis for analysis in the pre-notification group, and 42 in the no pre-notification group. Analysis was conducted on an intention-to-treat basis, and included all papers assigned to each intervention.

### Outcomes and estimation

At the end of follow up, 14/33 (42.4%) authors in the pre-notification condition had returned responses to the questionnaire, and 18/42 (42.9%) had returned response responses to the questionnaire in the no pre-notification condition.

The difference between the two arms is therefore -0.4% (95% CI -23.0% to 22.1%). A t-test found little evidence to support rejecting the null hypothesis of no difference between the two proportions, t (68.4) = -0.038, p = .968.

## Discussion

This randomised trail examined the effect sending authors of other randomised experiments examining the same question and included in a systematic review, but were being contacted for further information. The trail results imply that pre-notification does not improve response rates.

### Limitations

This study has several limitations. Firstly, the width of the 95% confidence intervals for the difference in the response rates is very large. This implies that the null result could be due to low precision, despite the point estimate being very close to the null value. The lack of precision could have been reduced by having a larger sample size, although this was caped due to the pragmatic nature of the inclusion criteria, or by having a more balanced randomisation list.

A second potential limitation is that is that the intervention used was the same intervention as the included study author’s studies had examined. This may have meant that contacted authors guessed that they were in the intervention or control arm of a randomised control trail examining the effect of pre-notification. This occurred for certain in one instance in the intervention arm. If so, then some degree of unblinding would have occurred, which might have biased the results. However, although ultimately unknowable, it seems probable that this would have only occurred for a minority of authors, in which case any bias is likely to be small.

Finally, there is a potential risk of bias due to study personnel being conducted unblinded. However, because most communication with the participants, prior to responses, was pre-written the magnitude of any bias this could introduce should be small.

### Interpretation

There is an extensive literature examining the role of pre-notification on response rates. This has generally found that pre-notification is beneficial to response, as summarised in, e.g., Edwards et al.[4]. This is contrary to the results of this study, which did not find evidence for an effect of pre-notification on response rates. This result is also contrary to the overall finding of the update to this review, in which this study was nested.

This could be for three reasons. Firstly, the true estimate might be lower than is typically thought. After removing studies at high or unclear risk of bias, we found that the ratio of the odds of responding given a pre-notification or no pre-notification decreased from OR = 1.38 (95%CI: 1.25-1.53) to OR = 1.11 (95% CI: 1.01-1.21) [5].

Secondly, the result of this study might be due to the small sample size, and thus low power. This is supported by the large confidence intervals for the risk difference. Any issue with power would be exacerbated if the effect estimate is smaller than the one typically used in the literature.

### Generalisability

Both reviews found substantive heterogeneity across studies. It is possible that some of this heterogeneity may be due to the effect of pre-notification differing depending on the context or population in which it is used. If this the case, then the studies results might not generalise to other context or study populations. Likewise, differences in results might different depending on the nature of the pre-notification (e.g. delay between sending of pre-notification and questionnaire, method of sending prenotification/questionnaire, etc). Either of these possibilities would limit the generalisability of this studies results.

## Conclusion

This randomised control trail sought to assess the impact of pre-notification on response to a request for more feedback by study authors, whose studies had been included in a systematic review. The study did not find evidence to support a difference in response across the control and intervention group. This is probably due to either the small sample size, or the effect of pre-notification being smaller than what it is typically thought.

## Data Availability

Data can be made available by contacting the primary author

## Other information

### Registration

This trial is not registered with any trail registry. However, the protocol was posted in advance on the Open Science Framework website.

### Protocol

The full trail protocol is available on the Open Science Framework website: https://osf.io/msv2w/

### Ethics

This study received ethical approval from the University of Bristol Life Sciences research ethics committee in December 2018 (Ref no. 75602).

