## Supplemental Table 1 for "Does pre-notification increase questionnaire response rates: a nested randomised control trial"

**Supplementary Table 1:** Pre-written communication

Pre-notification contact

Email title: Request for further information.

Dear [insert name]

We emailed you yesterday about your [insert date] paper '[insert title]', which has been included in a partial update to our 2009 Cochrane systematic review into improving response rates to questionnaires. If you are not too busy, we would be very grateful if you could answer the attached questions about the research methods you used?

Thank you for taking the time read this email, we look forwards to hearing back from you soon,

All the best,

Phil Edwards and Benji Woolf

In cases in which the study authors replied to the pre-notification email, they were sent the following communication:

Dear [insert name]

Thank you very much for your willingness to help us in our review. I have attached the survey to this email. If you are not too busy, we would be very grateful if you could answer the attached questions about the research methods you used. I hope you have a wonderful time on vacation!

All the best, and thank you again,

Benji

Follow up

Dear [insert name]

We emailed you last week with some questions about your [insert date] paper '[insert title]', which has been included in a partial update to our 2009 Cochrane systematic review into improving response rates to questionnaires. If you are not too busy, we would be very grateful if you could answer the attached questions about the research methods you used?

Thank you for taking the time read this email, we look forwards to hearing back from you soon,

All the best,

Phil Edwards and Benji Woolf

pre-notification

Dear [insert name]

Thank you for taking the time to read this email.  We are currently trying to update part of our 2009 Cochrane systematic review into improving response rates to questionnaires, and your [insert date] paper '[insert title]' was selected for inclusion. However, we were hoping you could provide us with some extra information about the methods you used, and would not mind answering some quick questions we will be emailing you tomorrow.

Thank you again for taking the time read this email, we look forwards to hearing back from you soon,

All the best,

 Phil Edwards and Benji Woolf

Non prenotification contact

Dear [insert name]

Thank you for taking the time to read this email.  We are currently trying to update part of our 2009 Cochrane systematic review into improving response rates to questionnaires, and your [insert date] paper '[insert title]' was selected for inclusion. If you are not too busy, we would be very grateful if you could answer the attached questions about the research methods you used?

Thank you again for taking the time read this email, we look forwards to hearing back from you soon,

All the best,

 Phil Edwards and Benji Woolf
