## Supplemental Table 2 for "Does pre-notification increase questionnaire response rates: a nested randomised control trial"

Supplementary Table 2: Questionnaire:

In the following boxed please could give further details about the methods you used to:

| The method used for random sequence generation |
| --- |
| The method used for allocation concealment |
| How participants were blinded/masked |
| How personnel were blinded/masked |
| How outcome assessors were blinded/masked |
| The delay between the administration of the pre-notification and the questionnaire |
| If you have conducted any other research addressing this question |
